# Longitudinal Prediction of Adolescent Depression from Environmental and Polygenic Risk Scores

**DOI:** 10.1101/2025.07.08.25331098

**Authors:** Eileen Y. Xu, Poppy Z. Grimes, Alex S.F. Kwong, Stephen M. Lawrie, Heather C. Whalley

**Author notes:** **Corresponding author:** Eileen Y. Xu; Division of Psychiatry, The Chancellor’s Building, 49 Little France Crescent, Edinburgh EH16 4SB.

## Abstract

Understanding adolescent depression risk is vital for mitigating its long-term adverse effects. Though polygenic risk scores (PRS) explain increasing proportions of heritable risk, environmental risk remains challenging to quantify, hindering prediction. Existing prediction models often examine environmental risk in isolation, and vary in the number of predictors used – ranging from 8 to 800+ variables, limiting generalisability. Here, we develop a model predicting adolescent depression symptoms (depRS) from a review of key environmental risk factors and assess depRS and PRS prediction of lifetime depression at 2-year follow-up. Using data from the Adolescent Brain Cognitive Development study (N=7 029), we generated PRS in European, African, American Admixed and East Asian ancestries from a recent trans-ancestry genome-wide study of major depression. We trained depRS using Elastic Net regression with 10-fold cross-validation to predict follow-up depression symptoms (age 11-13 years) from 23 baseline predictors (age 9-11 years), identified from systematic reviews with meta-analyses of risk factors. Parental depression, abuse, sleep duration and dieting emerged as top predictors of depression symptoms; depRS explained 16.9% of overall variance. depRS showed better-than-chance classification of parent-reported (AUC=0.68; 95% CI 0.63-0.72) lifetime depression at follow-up, associating with greater depression odds (OR=1.73; 95% CI: 1.57-1.91) than PRS (OR=1.42; 95% CI: 1.25-1.62). Combining depRS and PRS maximised accuracy (AUC=0.70; 95% CI 0.65-0.78). Though external validation of depRS across geographically and gender diverse cohorts is needed to assess generalisability, findings highlight sleep and dieting as potential targets for mitigating risk and demonstrate the utility of genetic scores in models predicting adolescent depression.

## Introduction

Adolescence is a key developmental period characterized by continued brain and bodily development alongside significant life transitions and growing independence. The incidence of depression also rises during adolescence; 47% of people with depressive disorders begin experiencing symptoms by age 25 [1, 2]. Depression during adolescence is associated with poorer health, educational attainment and life satisfaction in adulthood, particularly with younger age of onset [3–5]. Such research highlights the need for prevention and early intervention in at-risk youth, and the need for effective methods of quantifying depression risk.

Depression, like other mental health disorders, is highly heritable [6] and polygenic, with many hundreds of genetic variations contributing small amounts to overall risk [6–8]. However, genetic risk alone is likely to always be insufficient for accurate prediction given the complexity and heterogenous nature of depression [7, 9]; current polygenic risk scores (PRS) representing cumulative genetic risk explain ∼6% of depression variability in adult depression [8]. Additional sources of variance include environmental factors – the individual, family, peer, community and population-level influences on a young person’s life – and gene-environment interaction effects [5]. Unlike genetic variants, environmental factors can be modifiable; models predicting depression from environmental factors have the added benefit of highlighting opportunities to reduce this risk.

Existing prediction models show substantial variation in the number and complexity of feature sets used to predict depression. Common predictors across models include preceding depression symptoms [10–14], parental depression [10, 12, 14–16], stressful life events [10, 13, 16–19] and peer problems such as bullying [11, 12, 14, 15, 18, 19]. Some models also leverage genetic [17], brain [11, 20, 21]and biomarker [14, 20] data from large longitudinal studies, including as many as 885 features [14] and as few as 8 [10], with sample sizes for discovery data sets ranging from N<400 [10, 20] to N>8 000 [12, 14]. Feature sets across existing prediction models have also been determined *a priori*, based on factors such as data availability in discovery samples, clinical judgement and ease of measurement in addition to evidence from the literature [10, 18, 20]. As large cohort studies – required for sufficient statistical power – frequently differ in data availability, questionnaire use and study design, identifying common risk factors and targets for early intervention across prediction models can be challenging.

In this work, we aimed to (i) develop a prediction model using environmental factors measured at baseline (depRS) to estimate concurrent and 2-year follow up parent-reported depression symptoms (as a continuous measure). Additionally, we aimed to (ii) assess if our depRS could predict binary outcomes of self- and parent-reported lifetime Major Depressive Disorder (MDD) at a 2-year follow-up, and (iii) compare prediction of lifetime MDD using our depRS, a MDD PRS and their combined interaction. The depRS model was built in two stages, beginning with a review of key environmental risk factors associated with adolescent depression to develop a list of candidate features (Figure 1A). Parent-reported depression symptoms at 2-year follow-up were then regressed on candidate features which were present in baseline data from the Adolescent Brain Cognitive Development (ABCD) study [22, 23] using Elastic Net regression models (Figure 1B). Coefficients from this model were compared to a secondary Elastic Net with concurrent depression symptoms as the outcome to examine individual features and their relative importance to later depression. The review and Elastic Net modelling were not hypothesis-driven; for the latter analyses of depRS and PRS we hypothesized that (1) depRS prediction would outperform PRS, and (2) prediction using both depRS and PRS would explain additional variance in both self- and parent-reported lifetime MDD.

**Figure 1:**
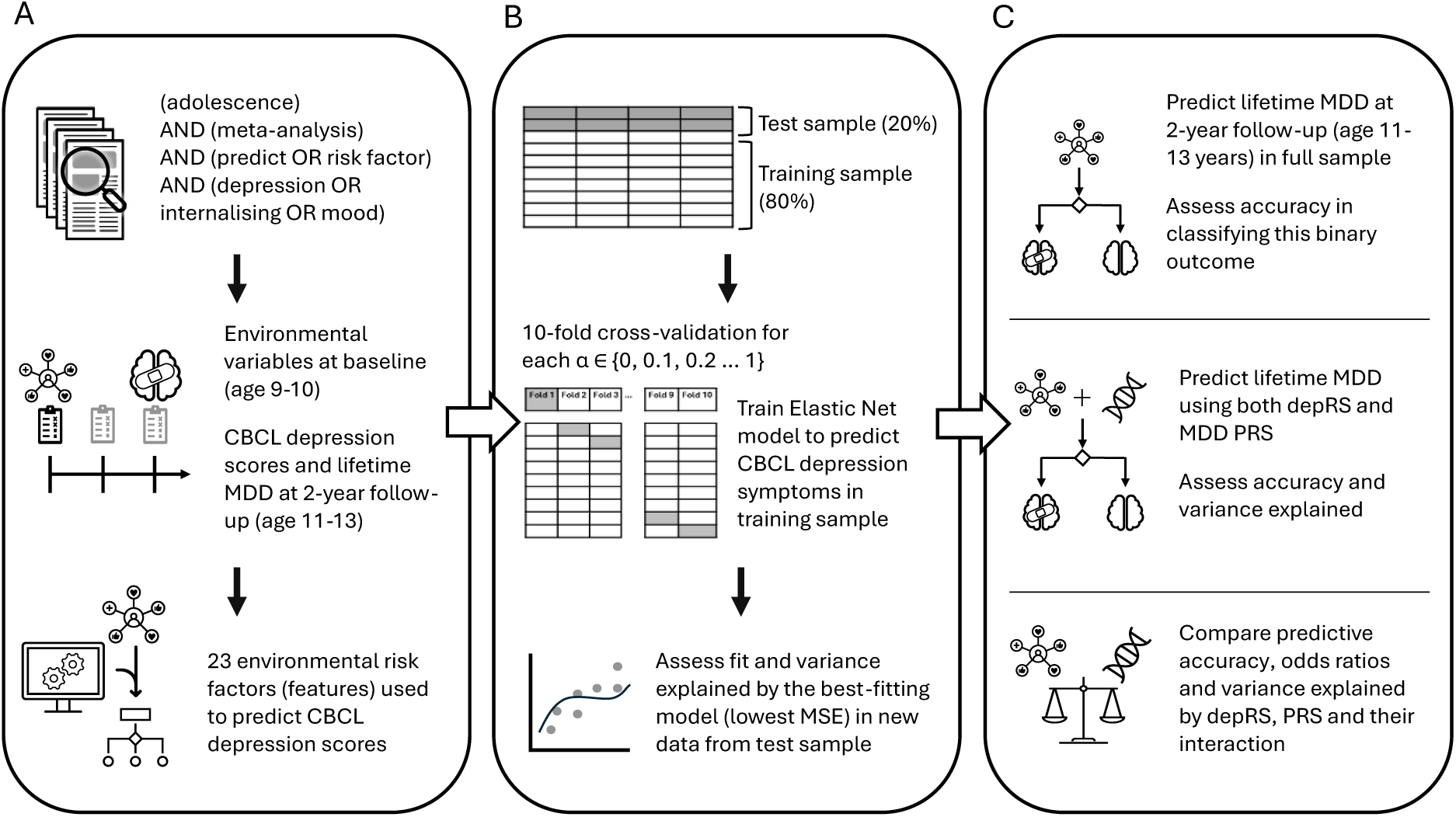
depRS development, training and evaluation process. (A) Risk factors were identified by reviewing environmental factors associated with depression from published review articles and meta-analyses. Once identified, these were matched to available data from ABCD study baseline assessments. (B) ABCD data were divided into training and test samples. Elastic net models were trained to predict depression scores at 2-year follow-up assessments using 10-fold cross-validation in the training sample. Coefficients from the best-fitting result were applied to unseen data in the test sample. (C) depRS and depression PRS were used to predict lifetime MDD at 2-year follow-up. Performance metrics from depRS-only, PRS-only and combined models were compared. ABCD = Adolescent Brain Cognitive Development study; CBCL = Child Behavior Checklist; MSE = Mean Squared Error; MDD = Major Depressive Disorder; depRS = environmental risk score for depression; PRS = Polygenic risk score for depression.

## Methods

### Risk factor review

MEDLINE, EMBASE and Web of Science Core Collection were searched for meta-analyses of risk factors for adolescent depression and/or internalising problems. As the objective was to identify established risk factors for inclusion in a feature-reduction machine learning analysis, grey literature and citation-matching searches were omitted. We included the following search terms: (adolescence) AND (meta-analysis) AND (predict OR risk factor) AND (depression OR internalising OR internalizing OR mood). Filters were included to limit results to meta-analyses and systematic reviews published in English between 1^st^ January 2000 and 1^st^ October 2024.

Articles reviewing interventions, treatments or therapies were removed, as well as any reviews which focused on the COVID-19 pandemic. Reviews specific to clinical populations/illnesses or single demographic sub-groups were also omitted. Of the 948 unique articles identified, 86 articles were selected following title and abstract screening. Full-text screening yielded a total 31 articles describing 38 candidate features with statistically significant associations with depression.

### ABCD Sample

The Adolescent Brain Cognitive Development (ABCD) study is a longitudinal cohort study of 11,880 children recruited from schools across 21 study sites, geographically distributed over the United States (abcdstudy.org). Detailed descriptions of ABCD design, recruitment and data collection protocols have been published elsewhere [22, 24, 25]. Briefly, eligible children were aged 9-10 years at baseline assessment with parents fluent in English or Spanish; children with limited English proficiency or severe sensory/neurological/medical issues which could affect the validity of collected data were excluded. ABCD study protocols were approved by a central Institutional Review Board (IRB) at the University of California, San Diego and sample recruitment and data collection was conducted in accordance with each site’s local IRB [25]. Parent/guardian informed consent and child assent was obtained at each assessment. Ethical approval for secondary data analysis using the ABCD dataset was obtained from the University of Edinburgh.

Data from baseline (age 9-10) and 2-year follow-up (age 11-13) assessments were included in the present analyses. Participants with missing predictor data were excluded from analyses; participants with missing outcome data were only excluded from the corresponding analysis (Table S1). One participant was randomly sampled from each family to obtain a sample of unrelated participants. Sample sizes and demographic characteristics for each analysis are presented in Table 1; see Table S2 for sample demographic characteristics by lifetime MDD status.

**Table 1:** Sample demographic characteristics for depRS and PRS analyses. Sample sizes and demographic characteristics for ABCD participants included in Elastic Net models predicting (i) concurrent and (ii) 2-year follow-up depression; and for participants included in depRS and PRS prediction of (iii) youth self-reported and (iv) parent-reported lifetime MDD at 2-year follow-up. PRS = polygenic risk score; NA = not applicable in this sample; GED = General Educational Development diploma; MDD = major depressive disorder. ^1^ To protect the privacy of study participants, demographic groups with fewer than 5 participants are denoted as <5 (<0.1%); numbers will sum to greater than the total sample size. ^2^ The 5-level race/ethnicity variable was coded in ABCD Releases from caregiver responses to “What race do you consider the child to be? Please check all that apply.” and “Do you consider the child Hispanic/Latino/Latina? (Yes/No)”. “Other race/ethnicity” consists of non-Hispanic youth where “Other race” or multiple of the following races/ethnicities were selected: White, Black/African American, Alaska Native, Native Hawaiian, Guamanian, Samoan, Other Pacific Islander, Asian Indian, Chinese, Filipino, Japanese, Korean, Vietnamese, Other Asian, Other Race, Refuse to Answer, Don’t Know. ^3^ Total income before taxes or deductions from all sources – e.g. wages, benefits, rent from properties, social security, child payments etc.

|  | (i) Baseline (N = 7,569) | (ii) 2-year follow-up (N = 7,029) | (iii) PRS youth (N = 6,299) | (iv) PRS parent (N = 6,259) |
| --- | --- | --- | --- | --- |
| Age, mean (SD), years |  |  |  |  |
| Baseline | 9.9 (0.6) | 9.9 (0.6) | 9.9 (0.6) | 9.9 (0.6) |
| 2-year follow-up | NA | 12 (0.7) | 12 (0.7) | 12 (0.7) |
| Gender identity <sup>1</sup>, n (%) |  |  |  |  |
| Cisgender male | 3,990 (52.7%) | 3,729 (53.1%) | 3,366 (53.4%) | 3,334 (53.3%) |
| Cisgender female | 3,575 (47.2%) | 3,296 (46.9%) | 2,930 (46.5%) | 2,922 (46.7%) |
| Transgender male | <5 (<0.1%) | <5 (<0.1%) | <5 (<0.1%) | <5 (<0.1%) |
| Transgender female | <5 (<0.1%) | <5 (<0.1%) | <5 (<0.1%) | <5 (<0.1%) |
| Assigned sex at birth, n (%) |  |  |  |  |
| Male | 3,994 (52.7%) | 3,734 (53.1%) | 3,370 (53.5%) | 3,338 (53.3%) |
| Female | 3,575 (47.3%) | 3,295 (46.9%) | 2,929 (46.5%) | 2,921 (46.7%) |
| Race/Ethnicity <sup>2</sup>, n (%) |  |  |  |  |
| Asian | 166 (2.2%) | 151 (2.1%) | 61 (1.0%) | 61 (1.0%) |
| Black | 962 (12.7%) | 845 (12.0%) | 794 (12.6%) | 784 (12.5%) |
| Hispanic | 1,405 (18.6%) | 1,273 (18.1%) | 1,129 (17.9%) | 1,119 (17.9%) |
| White | 4,206 (55.6%) | 3,992 (56.8%) | 3,778 (60.0%) | 3,764 (60.1%) |
| Other | 830 (11.0%) | 768 (10.9%) | 537 (8.5%) | 531 (8.5%) |
| Total household income p.a. <sup>3</sup>, n (%) |  |  |  |  |
| < \$5,000 | 239 (3.2%) | 202 (2.9%) | 175 (2.8%) | 172 (2.7%) |
| \$5,000 - \$11,999 | 279 (3.7%) | 236 (3.4%) | 210 (3.3%) | 207 (3.3%) |
| \$12,000 - \$15,999 | 178 (2.4%) | 152 (2.2%) | 134 (2.1%) | 133 (2.1%) |
| \$16,000 - \$24,999 | 330 (4.4%) | 293 (4.2%) | 257 (4.1%) | 255 (4.1%) |
| \$25,000 - \$34,999 | 443 (5.9%) | 404 (5.7%) | 362 (5.7%) | 361 (5.8%) |
| \$35,000 - \$49,999 | 651 (8.6%) | 593 (8.4%) | 545 (8.7%) | 538 (8.6%) |
| \$50,000 - \$74,999 | 1,065 (14.1%) | 981 (14.0%) | 904 (14.4%) | 900 (14.4%) |
| \$75,000 - \$99,999 | 1,126 (14.9%) | 1,080 (15.4%) | 974 (15.5%) | 969 (15.5%) |
| \$100,000 - \$199,999 | 2,368 (31.3%) | 2,252 (32.0%) | 2,027 (32.2%) | 2,018 (32.2%) |
| <\$200,000 | 890 (11.8%) | 836 (11.9%) | 711 (11.3%) | 706 (11.3%) |
| Parental education, n (%) |  |  |  |  |
| <High school | 337 (4.5%) | 282 (4.0%) | 235 (3.7%) | 229 (3.7%) |
| High school/GED | 718 (9.5%) | 641 (9.1%) | 577 (9.2%) | 573 (9.2%) |
| Some College/Associate degree | 2,157 (28.5%) | 1,963 (27.9%) | 1,808 (28.7%) | 1,794 (28.7%) |
| Bachelor's degree | 2,242 (29.6%) | 2,132 (30.3%) | 1,941 (30.8%) | 1,935 (30.9%) |
| Postgraduate degree | 2,115 (27.9%) | 2,011 (28.6%) | 1,738 (27.6%) | 1,728 (27.6%) |
| Lifetime MDD, n (%) |  |  |  |  |
| 0 | NA | NA | 6,088 (96.7%) | 6,011 (96.0%) |
| 1 | NA | NA | 211 (3.3%) | 248 (4.0%) |

### Depression measures

Elastic Net models were trained on standardised Child Behaviour Checklist (CBCL) [26] depression subscale scores collected at baseline and 2-year follow-up assessments. The CBCL provides parent-reported measures of dimensional psychopathology and is widely used in children aged 6-18 years, with clinically validated [27] subscales corresponding to Diagnostic and Statistical Manual of Mental Disorders (DSM) symptom criteria for major depression [26, 27].

Lifetime MDD was assessed at 2-year follow-up using the computerized Kiddie Schedule for Affective Disorders and Schizophrenia, version 5 (KSADS-COMP) [28], combining past and current (past 2 weeks) MDD diagnoses. As discrepancies between parent- and child-reports of depression are common in adolescence [29–31] and have been reported previously in ABCD [30, 32], we conducted separate analyses for parent- and child-reports of lifetime MDD.

### Predictors (features)

We matched candidate features to existing variables in ABCD baseline data, excluding features with >20% missing data and using summary scores where available. This yielded a total 23 features measured at baseline; 32 features were available at 2-year follow-up. In this study, we defined the risk factor “gender” [33] as *current gender identity* (parent-reported), where “male” includes both cisgender and transgender men (boys) and “female” includes both cisgender and transgender women (girls). A full list of risk factors and corresponding ABCD variables is available in Table S3.

### PRS

Multi-ancestry and European-only GWAS summary statistics were obtained from the MDD Working Group of the Psychiatric Genomics Consortium [8]. Using the 1000 Genomes reference panel superpopulations [34], we calculated the top 10 genetic principal components (PCs) and used the first 6 PCs to predict the genetic clusters into European, African, American Admixed (Hispanic/Latino), and East Asian ancestries. PRS were generated in all ancestries using multi-ancestry summary statistics; sensitivity analysis using European-only summary statistics in European ancestries is reported in the Supplementary Information.

We developed PRS using SBayesRC (in GCTB v2.5.2) [35], a method which uses functional genomic annotations to improve polygenic prediction in diverse ancestries [36]. We used ancestry-specific linkage disequilibrium reference panels from UK Biobank and the 7M SNP functional annotations derived from S-LDSC BaselineLDv2.2 [37]. An ancestry-specific reference panel was not available for American Admixed ancestry; given the high levels of European ancestry in this population [38], the European reference panel was used as a proxy. Finally, PRS based on the SBayesRC SNP predictions output were generated using PLINK v1.9 [39].

### Statistical analysis

Elastic Net regression models to predict concurrent and follow-up depression scores from baseline features were trained using the ‘glmnet’ package (v4.1.8) [40] in R (v4.4.3) [41]. Elastic Net hyperparameters were tuned using 10-fold cross-validation in the training (80%) sample. Coefficients from the best-fitting (lowest mean-squared error; MSE) model were then used to weight predictor variables in the test (20%) sample for internal validation. Further details on hyperparameter tuning are provided in the Supplementary Information alongside a brief description of supervised machine learning and the Elastic Net algorithm.

We examined depRS and PRS prediction of lifetime MDD (Figure 1C) through step-wise comparisons of (1) depRS-only or PRS-only (2) depRS + PRS (3) depRS*PRS logistic regression models using Likelihood Ratio Tests (α = .05). PRS models also included the first 5 genetic PCs as covariates. We report odds ratios (ORs) for depRS, PRS and their interaction; overall fit and classification of lifetime MDD status are reported as Tjur’s R^2^ and area under the receiver operating curve (AUC; values > 0.5 indicate better-than-chance performance), respectively.

### Sensitivity analysis

To address potential differences in feature importance between gender identity and sex assigned at birth, we conducted a sensitivity analysis using participants’ assigned sex at birth in place of gender identity to predict depression scores. As depression, anxiety and emotional problems are highly correlated in childhood and adolescence [5], a parallel analysis was conducted to predict CBCL internalising subscale scores. To assess additional candidate features missing from baseline data, we repeated the cross-sectional depRS analysis in 2-year follow-up data using 32 available features; insufficient follow-up data at the time of analysis precluded longitudinal analyses. Finally, we conducted a sensitivity analysis predicting lifetime MDD using European GWAS summary statistics for PRS in participants of European ancestry.

## Results

### Risk factor review

We identified 38 risk factors for adolescent depression from 31 review articles. Individual-level risk factors included gender [33, 42–46] and existing mental health issues^48^. Additional individual-level risk factor categories were substance use [42, 47], adverse experiences [48–50], cognitive skills [42, 51–55], lifestyle factors [42, 45, 47, 56–60] and technology use [42, 44, 46, 61] (Table 2). Microsystem-level factors included bullying [43, 62–65], family environment [66–69] and socioeconomic status [50, 70]; community safety and discrimination [71] were the only macrosystem-level factors identified.

**Table 2:**
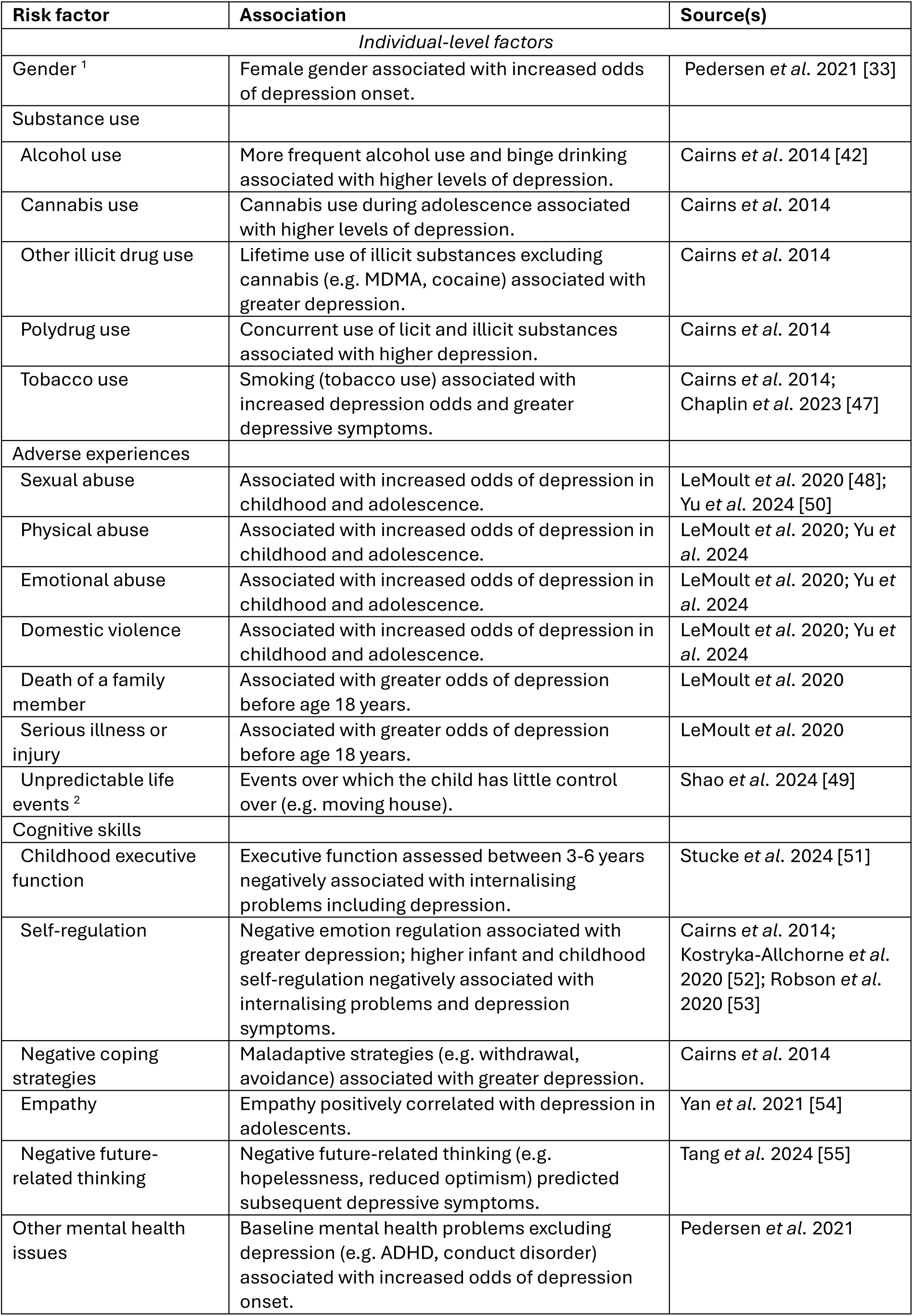
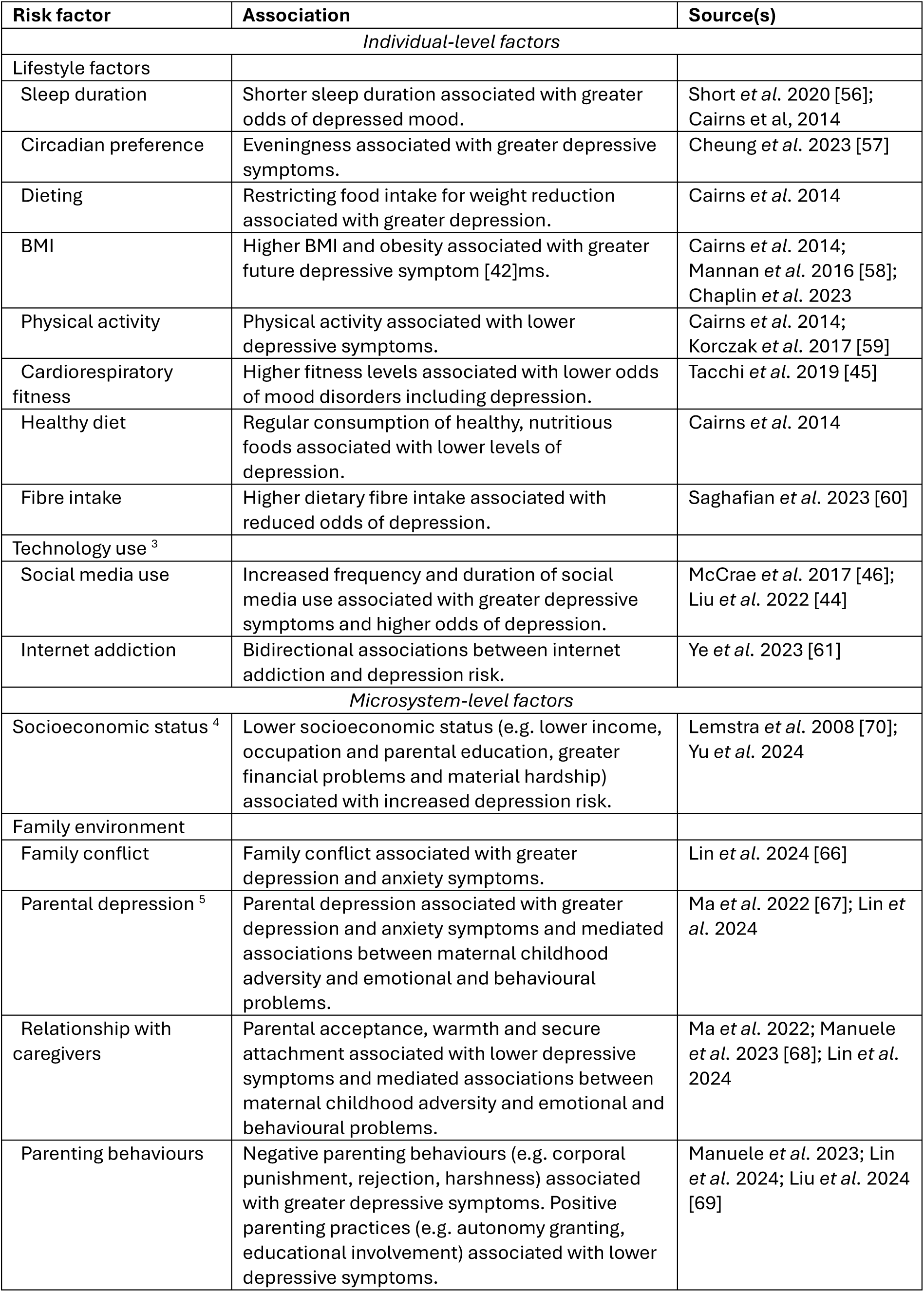
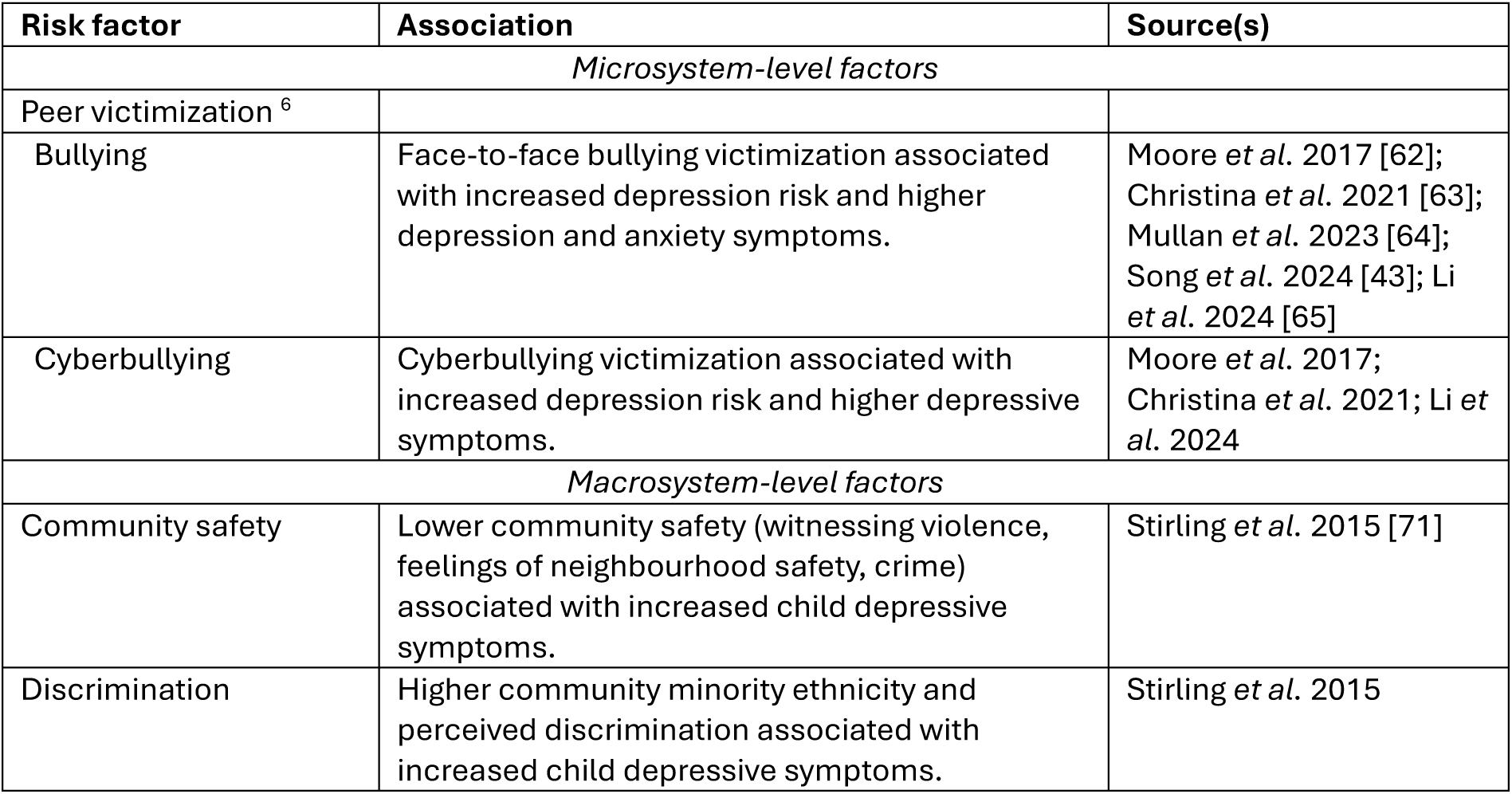
Risk factors associated with depression. Risk factors associated with depression in adolescence, grouped by socioecological model level. ^1^ Gender was also identified as a significant moderating effect in Cairns *et al.* 2014 [42], Liu *et al.* 2022 [44], Song *et al.* 2024 [43], Tacchi *et al.* 2019 [45] and McCrae *et al.* 2017 [46]. No reviews distinguished between gender identity and assigned sex at birth due to a lack of reporting in the original studies. ^2^ Shao *et al.* 2024 [49] examined a single construct termed “early life uncertainty” –defined as experiences over which the child had little to no control over – in relation to internalising problems. This overlapped with adverse experiences described by LeMoult *et al.* 2020 [48] and Yu *et al.* 2024 [50] and included parental job loss, parental separation or divorce, residential mobility and changes in family structure. ^3^ Cairns *et al.* 2014 [42] examined “media use” as a broad construct which included television, gaming and internet use. ^4^ LeMoult *et al.* 2020 [48] found no association between poverty and depression odds over 9 studies; they noted significant heterogeneity across effect sizes. ^5^ Pedersen *et al.* 2023 [33] found no association between parental history of depression and child depression onset over 3 studies; they reported moderate-to-high heterogeneity across risk factors. ^6^ Moore *et al.* 2017 [62] included both face-to-face bullying and cyberbullying in their meta-analysis of bullying victimization.

| Risk factor | Association | Source(s) |
| --- | --- | --- |
| *Individual-level factors* |  |  |
| Gender <sup>1</sup> | Female gender associated with increased odds of depression onset. | Pedersen *et al.* 2021 [33] |
| Substance use |  |  |
| Alcohol use | More frequent alcohol use and binge drinking associated with higher levels of depression. | Cairns *et al.* 2014 [42] |
| Cannabis use | Cannabis use during adolescence associated with higher levels of depression. | Cairns *et al.* 2014 |
| Other illicit drug use | Lifetime use of illicit substances excluding cannabis (e.g. MDMA, cocaine) associated with greater depression. | Cairns *et al.* 2014 |
| Polydrug use | Concurrent use of licit and illicit substances associated with higher depression. | Cairns *et al.* 2014 |
| Tobacco use | Smoking (tobacco use) associated with increased depression odds and greater depressive symptoms. | Cairns *et al.* 2014; Chaplin *et al.* 2023 [47] |
| Adverse experiences |  |  |
| Sexual abuse | Associated with increased odds of depression in childhood and adolescence. | LeMoult *et al.* 2020 [48]; Yu *et al.* 2024 [50] |
| Physical abuse | Associated with increased odds of depression in childhood and adolescence. | LeMoult *et al.* 2020; Yu *et al.* 2024 |
| Emotional abuse | Associated with increased odds of depression in childhood and adolescence. | LeMoult *et al.* 2020; Yu *et al.* 2024 |
| Domestic violence | Associated with increased odds of depression in childhood and adolescence. | LeMoult *et al.* 2020; Yu *et al.* 2024 |
| Death of a family member | Associated with greater odds of depression before age 18 years. | LeMoult *et al.* 2020 |
| Serious illness or injury | Associated with greater odds of depression before age 18 years. | LeMoult *et al.* 2020 |
| Unpredictable life events <sup>2</sup> | Events over which the child has little control over (e.g. moving house). | Shao *et al.* 2024 [49] |
| Cognitive skills |  |  |
| Childhood executive function | Executive function assessed between 3-6 years negatively associated with internalising problems including depression. | Stucke *et al.* 2024 [51] |
| Self-regulation | Negative emotion regulation associated with greater depression; higher infant and childhood self-regulation negatively associated with internalising problems and depression symptoms. | Cairns *et al.* 2014; Kostryka-Allchorne *et al.* 2020 [52]; Robson *et al.* 2020 [53] |
| Negative coping strategies | Maladaptive strategies (e.g. withdrawal, avoidance) associated with greater depression. | Cairns *et al.* 2014 |
| Empathy | Empathy positively correlated with depression in adolescents. | Yan *et al.* 2021 [54] |
| Negative future-related thinking | Negative future-related thinking (e.g. hopelessness, reduced optimism) predicted subsequent depressive symptoms. | Tang *et al.* 2024 [55] |
| Other mental health issues | Baseline mental health problems excluding depression (e.g. ADHD, conduct disorder) associated with increased odds of depression onset. | Pedersen *et al.* 2021 |
| *Individual-level factors* |  |  |
| Lifestyle factors |  |  |
| Sleep duration | Shorter sleep duration associated with greater odds of depressed mood. | Short *et al.* 2020 [56]; Cairns *et al.* 2014 |
| Circadian preference | Eveningness associated with greater depressive symptoms. | Cheung *et al.* 2023 [57] |
| Dieting | Restricting food intake for weight reduction associated with greater depression. | Cairns *et al.* 2014 |
| BMI | Higher BMI and obesity associated with greater future depressive symptom [42]ms. | Cairns *et al.* 2014; Mannan *et al.* 2016 [58]; Chaplin *et al.* 2023 |
| Physical activity | Physical activity associated with lower depressive symptoms. | Cairns *et al.* 2014; Korczak *et al.* 2017 [59] |
| Cardiorespiratory fitness | Higher fitness levels associated with lower odds of mood disorders including depression. | Tacchi *et al.* 2019 [45] |
| Healthy diet | Regular consumption of healthy, nutritious foods associated with lower levels of depression. | Cairns *et al.* 2014 |
| Fibre intake | Higher dietary fibre intake associated with reduced odds of depression. | Saghafian *et al.* 2023 [60] |
| Technology use <sup>3</sup> |  |  |
| Social media use | Increased frequency and duration of social media use associated with greater depressive symptoms and higher odds of depression. | McCrae *et al.* 2017 [46]; Liu *et al.* 2022 [44] |
| Internet addiction | Bidirectional associations between internet addiction and depression risk. | Ye *et al.* 2023 [61] |
| *Microsystem-level factors* |  |  |
| Socioeconomic status <sup>4</sup> | Lower socioeconomic status (e.g. lower income, occupation and parental education, greater financial problems and material hardship) associated with increased depression risk. | Lemstra *et al.* 2008 [70]; Yu *et al.* 2024 |
| Family environment |  |  |
| Family conflict | Family conflict associated with greater depression and anxiety symptoms. | Lin *et al.* 2024 [66] |
| Parental depression <sup>5</sup> | Parental depression associated with greater depression and anxiety symptoms and mediated associations between maternal childhood adversity and emotional and behavioural problems. | Ma *et al.* 2022 [67]; Lin *et al.* 2024 |
| Relationship with caregivers | Parental acceptance, warmth and secure attachment associated with lower depressive symptoms and mediated associations between maternal childhood adversity and emotional and behavioural problems. | Ma *et al.* 2022; Manuele *et al.* 2023 [68]; Lin *et al.* 2024 |
| Parenting behaviours | Negative parenting behaviours (e.g. corporal punishment, rejection, harshness) associated with greater depressive symptoms. Positive parenting practices (e.g. autonomy granting, educational involvement) associated with lower depressive symptoms. | Manuele *et al.* 2023; Lin *et al.* 2024; Liu *et al.* 2024 [69] |
| *Microsystem-level factors* |  |  |
| Peer victimization <sup>6</sup> |  |  |
| Bullying | Face-to-face bullying victimization associated with increased depression risk and higher depression and anxiety symptoms. | Moore *et al.* 2017 [62]; Christina *et al.* 2021 [63]; Mullan *et al.* 2023 [64]; Song *et al.* 2024 [43]; Li *et al.* 2024 [65] |
| Cyberbullying | Cyberbullying victimization associated with increased depression risk and higher depressive symptoms. | Moore *et al.* 2017; Christina *et al.* 2021; Li *et al.* 2024 |
| *Macrosystem-level factors* |  |  |
| Community safety | Lower community safety (witnessing violence, feelings of neighbourhood safety, crime) associated with increased child depressive symptoms. | Stirling *et al.* 2015 [71] |
| Discrimination | Higher community minority ethnicity and perceived discrimination associated with increased child depressive symptoms. | Stirling *et al.* 2015 |

### depRS coefficients

23 candidate features had corresponding questionnaire-based variables in ABCD baseline data (Table S3). The best-fitting depRS model predicting 2-year depression scores retained 22 out of 23 features, though the majority (16) had coefficients close to zero (Table S4). In the test sample, this model explained 16.9% variance in CBCL depression scores (MSE = 0.83). For CBCL depression scores measured concurrently with environmental factors, the best-fitting model explained 18.3% of depression score variance in the test sample (MSE = 0.82). Traumatic experiences, parental depression, sleep duration and dieting were leading contributors to depression at both timepoints (Figure 2A; Table S4) – coefficients were similar across timepoints for parental depression (concurrent β = 0.36; Y2 β = 0.30) and witnessing domestic violence (concurrent β = 0.19; Y2 β = 0.17).

**Figure 2:**
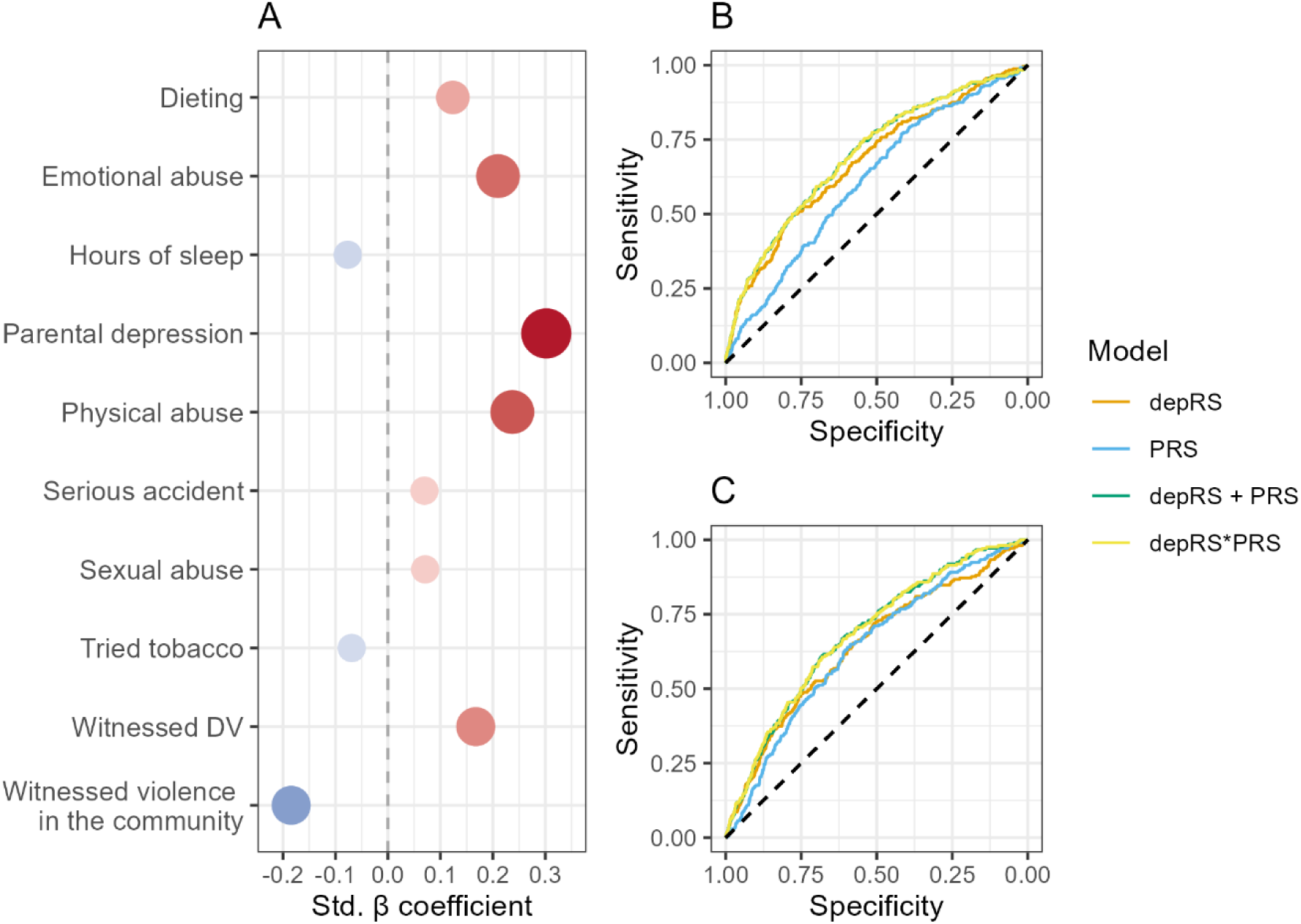
Top 10 depRS features and prediction of lifetime MDD. (A) Top 10 depRS features associated with CBCL depression scores at 2-year follow-up; point sizes represent absolute value and are coloured based on direction. (B) ROC curves of depRS and PRS classification of parent-reported and (C) youth self-reported lifetime MDD at follow-up. The dashed diagonal line represents a model with predictive accuracy which is no better than chance (AUC of 0.5); models with greater accuracy have curves which lie closer to the top-left corner of the ROC plot. CBCL = Child Behavior Checklist; DSM5 = Diagnostic and Statistical Manual for Mental Disorders 5^th^ Edition; depRS = environmental risk score for depression. AUC = Area Under the Curve; MDD = Major Depressive Disorder; PRS = Polygenic risk score for depression; ROC = Receiver Operating Curve.

### depRS and PRS predicting lifetime MDD

depRS was associated with greater odds of parent-reported lifetime MDD (OR = 1.73 [95% CI 1.57 – 1.91]), and to a lesser extent, youth self-reported MDD (OR = 1.53 [95% CI 1.37 – 1.71]). depRS alone classified lifetime MDD status with better-than-chance accuracy (Parent: AUC = 0.68 [95% CI 0.63 – 0.72]; Youth: AUC = 0.64 [95% CI 0.59 – 0.68]), outperforming PRS-only prediction for both outcomes (Figure 2, Table 3). The linear combination of depRS and PRS improved overall model fit (Parent: χ^2^ = 23.76, p<.001; Youth: χ^2^ = 28.03, p<.001), explaining additional variance in lifetime MDD compared to either score alone (Table 3). This also improved prediction accuracy for self-reported, but not parent-reported, lifetime MDD (AUC = 0.67 [95% CI 0.62 – 0.71]). Including a depRS x PRS interaction term maximised classification of parent-reported lifetime MDD (AUC = 0.70 [95% CI 0.64 – 0.77]); however, this interaction did not significantly associate with lifetime MDD (Table 3).

**Table 3:** depRS and PRS prediction of lifetime MDD. Model fit and odds ratios from logistic regression models predicting youth self-reported and parent-reported lifetime MDD, assessed at 2-year follow-up. AUC = area under the receiver operator curve; RMSE = root mean square error; MDD = major depressive disorder; depRS = linear predictor from Elastic Net model predicting depression scores at 2-year follow-up from baseline environmental factors; PRS = polygenic risk score. NA = not estimated in this model. *p<0.05, **p<0.01, ***p<0.001

| Model | Model fit |  |  | Odds Ratio [95% CI] |  |  |
| --- | --- | --- | --- | --- | --- | --- |
|  | AUC [95% CI] | R<sup>2</sup> | RMSE | depRS | PRS | depRS * PRS |
| **Youth self-reported lifetime MDD** |  |  |  |  |  |  |
| **depRS only** | 0.64 [0.59 – 0.68] | 0.010 | 0.179 | 1.53 [1.37 – 1.71]*** | NA | NA |
| **PRS only** | 0.62 [0.53 – 0.67] | 0.006 | 0.180 | NA | 1.31 [1.15 – 1.50]*** | NA |
| **depRS + PRS** | 0.67 [0.62 – 0.71] | 0.015 | 0.179 | 1.48 [1.32 – 1.65]*** | 1.26 [1.10 – 1.45]*** | NA |
| **depRS \* PRS** | 0.67 [0.63 – 0.72] | 0.015 | 0.179 | 1.47 [1.30 – 1.64]*** | 1.24 [1.07 – 1.44]** | 1.03 [0.93 – 1.14] |
| **Parent-reported lifetime MDD** |  |  |  |  |  |  |
| **depRS only** | 0.68 [0.63 – 0.72] | 0.024 | 0.193 | 1.73 [1.57 – 1.91]*** | NA | NA |
| **PRS only** | 0.59 [0.56 – 0.60] | 0.005 | 0.195 | NA | 1.44 [1.26 – 1.65]*** | NA |
| **depRS + PRS** | 0.68 [0.60 – 0.74] | 0.028 | 0.192 | 1.71 [1.55 – 1.89]*** | 1.35 [1.18 – 1.54]*** | NA |
| **depRS \* PRS** | 0.70 [0.64 – 0.77] | 0.028 | 0.192 | 1.72 [1.55 – 1.91]*** | 1.37 [1.17 – 1.59]*** | 0.98 [0.89 – 1.09] |

### Sensitivity analysis

depRS analyses using sex assigned at birth had reversed coefficient directions for community safety, loss of a loved one and female sex (Table S4). depRS features explained greater variance in internalising scores compared to depression (1.7% - 2.5% increase); sexual abuse was also a stronger predictor of internalising scores (Table S4). Models predicting concurrent CBCL depression and internalising scores based on features at 2-year follow-up (Table S3) explained 8.6% - 13.7% greater variance compared to their corresponding baseline models (Table S6). Finally, combined depRS and PRS analyses using European-only summary statistics for individuals of European ancestry yielded comparable results (Table S7).

## Discussion

In this study, we developed a model predicting adolescent depression based on well-evidenced environmental risk factors (depRS), comparing its predictive accuracy against PRS from multiple ancestries. depRS was comprised of 23 environmental factors measured at age 9-10 years which together with the PRS, predicted parent-reported lifetime MDD at 2-year follow-up with 70% accuracy. depRS alone showed better-than-chance accuracy in classifying both youth self-reported (AUC = 0.64; R^2^ = 1.0%) and parent-reported (AUC = 0.68; R^2^ = 2.4%) MDD and associated with a greater increase in depression odds (OR = 1.53 – 1.73) compared to PRS (OR = 1.31 – 1.44). The feature set explained 16.9% of depression score variance at 2-year follow-up (age 11-13 years) and 18.3% cross-sectionally.

In contrast to previous models, we trained depRS on a dimensional measure of depression. While re-training depRS to predict a binary outcome would likely improve prediction accuracy, depRS did not perform poorly in comparison with existing prediction models, where AUC values can range from 0.64 – 0.79 [10, 11, 13, 14, 18–20]. Diagnostic models across other psychiatric traits such as schizophrenia and autism spectrum disorder show similar lower bounds [72], even after incorporating genetic variants [73]. The relatively small depRS feature set also explained a similar proportion of variance in cross-sectional depression scores (R^2^ = 0.18) compared to previous work in ABCD using nearly 400 variables including fMRI features (R^2^ = 0.16) [15]. Together, these findings suggest that more parsimonious prediction models with well-evidenced features can perform at least on par with more complex models – particularly if accuracy can be boosted by incorporating PRS.

Consistent with previous findings, caregiver depression and physical/emotional abuse were strong predictors of depression [5, 10, 50]. While experiences of trauma and adversity cannot be undone on an individual-level, these risk factors remain important for identifying young people at greater risk of depression; greater public awareness of the signs of abuse may also foster earlier intervention and help mitigate exposure on a broader scale. In terms of more modifiable risk factors, our results also highlighted shorter sleep duration and dieting as key predictors of future depression symptoms, consistent with observational epidemiological evidence [42, 56, 74].

The relationship between sleep and depression is well-documented and bidirectional, with disturbed sleep being both a symptom of and a contributor to worsening depression in adolescents [42, 56]. Thus, a focus on improving sleep quantity and quality may be beneficial in mitigating future depression risk. This aligns with previous priority-setting work involving young people and key stakeholders, which highlighted the benefits of targeting more accessible everyday experiences in early intervention [75]. While randomised controlled trials (RCTs) of universal sleep-based intervention programs – e.g. school-wide educational approaches – have shown mixed results thus far [76], selective interventions have improved perceived sleep quality and behavioural problems in at-risk adolescents [77]. Given the bidirectional associations between sleep and depression, additional work leveraging multimodal sleep data may be beneficial in improving sleep-based interventions for adolescent depression.

Previous work has identified associations between depression and dieting, however the nature of this relationship is less clear [42]. Dieting is highly prevalent in adolescent girls (∼50%) and increasingly in boys (∼25%) [78]; like depression, disordered eating behaviours can also persist into adulthood [78, 79]. This association may reflect a comorbidity with eating disorders, or point to shared mechanisms in the aetiology of both disorders in adolescence. Indeed, previous research has identified that depression symptoms and disordered eating behaviours develop concurrently in adolescence with shared alterations in brain structure observed prior to symptom onset [74]. Further investigation of underlying biological mechanisms is required to elucidate this relationship, with recent evidence suggesting a causal link between relative carbohydrate intake and lower depression risk [80].

### Strengths and limitations

Large feature sets of *a priori* predictor variables that are costly or difficult to routinely measure can limit the generalisability of prediction models to new samples, restricting their utility. Here, we present a more parsimonious feature set where candidate features were instead selected from an initial review of risk factors. This study included >6 000 participants with multi-ancestry PRS generated from the largest and most inclusive GWAS of depression to date [8], lending greater statistical power for genetic analyses that have often excluded participants of non-European ancestry [8, 9, 81]. Additionally, the ABCD sample allowed for the explicit definition of gender in this study, providing inclusivity and transparency in the reporting of sex as a risk factor [33]. However, due to the binary categorization of sex and gender in existing literature, we note that this work remains complicit in the marginalisation of gender non-conforming and intersex young people and emphasize the need for greater inclusion of these young people at increased risk of depression [5] in future longitudinal studies.

It is important to recognize that the generalisability of our depRS is limited by a lack of external validation. Though ABCD aimed to recruit a representative sample of US households [22], families with higher incomes and more years of education remain overrepresented [82], as is common in many large longitudinal studies. While challenging, validation across geographically diverse cohorts with greater inclusion of underrepresented young people from lower-income families remains necessary, as it is currently unclear whether these findings reflect the experiences of young people outside the US. Only one model from the existing literature – Identifying Depression Early in Adolescence Risk Score, trained in a Brazilian study [19] – has been externally validated, performing better-than-chance in Nepal [83], Nigeria [84], the US [85, 86] and the UK [19]. Additionally, as our depRS accuracy differed across parent- and youth self-reported depression outcomes, incorporating multi-informant measures may help evaluate the clinical relevance of these findings.

While we attempted to synthesise meta-analytic evidence on environmental risk factors for adolescent depression, the final depRS feature set is far from comprehensive. First, a number of community-level and structural factors – such as community connectedness [71], academic pressure [87] and societal expectations [87, 88] – were excluded, likely due to the narrow scope of our search. Lastly, several candidate features could not be examined longitudinally due to a lack of follow-up data; including these features in a future model may improve depRS prediction accuracy.

The longitudinal nature of the ABCD study provides ample opportunities for building on this work as participants navigate the rapidly changing landscape of psychosocial, physical and brain development in adolescence. While we primarily examined depression at a single time-point, the interplay of genetic and environmental factors in depression is not static [81], and depression symptoms may spontaneously remit, recur, persist into adulthood or signal the onset of disorders such as schizophrenia and bipolar disorder [5, 79]. Therefore, a trajectory-based or repeated measures approach where the contributions of environmental risk factors are integrated with genetic, neuroimaging and biomarker data may provide deeper insight into the temporal mechanisms underlying depression risk and its relation to longer-term health.

## Conclusions

Here, we demonstrate that depRS, a weighted sum of 23 risk factors for adolescent depression, predicts depression symptoms and lifetime MDD status at 2-year follow-up with better-than-chance accuracy (AUC: 65 – 68%). Compared to polygenic risk, depRS showed greater accuracy and associated with a greater increase in odds for parent- and self-reported MDD (OR: 1.53 – 1.73). No significant depRS-PRS interaction was found, however combining the two scores significantly improved model fit with a maximum AUC accuracy of 70%. Results highlighted parent/caregiver depression and abuse as key risk factors, identifying shorter sleep and dieting as possible targets for modifying risk. External validation across geographically and culturally diverse cohorts is required to evaluate the generalisability of these results, and future work integrating brain- and biomarker-based predictors may help boost prediction and uncover mechanisms underlying these associations. Finally, while depRS and other risk prediction models provide proof-of-concept, significant public and patient consultation is required to address the practical and ethical implications [89] of prediction modelling, particularly where mental health resources are scarce [90].

## Data Availability

ABCD Study data are available upon request https://abcdstudy.org/scientists/data-sharing/. Summary statistics used to generate PRS are available for public download from the Psychiatric Genomics Consortium https://pgc.unc.edu/for-researchers/download-results/. Functional genomics annotation files used to generate PRS are available for download from https://sbayes.pctgplots.cloud.edu.au/data/SBayesRC/resources/v2.0/Annotation/. All R code used in the present analyses are available on GitHub https://github.com/EileenYXu/envRS. 

https://pgc.unc.edu/for-researchers/download-results/

https://sbayes.pctgplots.cloud.edu.au/data/SBayesRC/resources/v2.0/Annotation/

https://github.com/EileenYXu/envRS

## Acknowledgements

Data used in the preparation of this article were obtained from the Adolescent Brain Cognitive Development^SM^ (ABCD) Study (https://abcdstudy.org), held in the NIMH Data Archive (NDA). This is a multisite, longitudinal study designed to recruit more than 10,000 children age 9-10 and follow them over 10 years into early adulthood. The ABCD Study® is supported by the National Institutes of Health and additional federal partners under award numbers U01DA041048, U01DA050989, U01DA051016, U01DA041022, U01DA051018, U01DA051037, U01DA050987, U01DA041174, U01DA041106, U01DA041117, U01DA041028, U01DA041134, U01DA050988, U01DA051039, U01DA041156, U01DA041025, U01DA041120, U01DA051038, U01DA041148, U01DA041093, U01DA041089, U24DA041123, U24DA041147. A full list of supporters is available at https://abcdstudy.org/federal-partners.html. A listing of participating sites and a complete listing of the study investigators can be found at https://abcdstudy.org/consortium_members/. ABCD consortium investigators designed and implemented the study and/or provided data but did not necessarily participate in the analysis or writing of this report. This manuscript reflects the views of the authors and may not reflect the opinions or views of the NIH or ABCD consortium investigators. The ABCD data repository grows and changes over time. The ABCD data used in this report came from NIMH Data Archive Digital Object Identifier (DOI): https://dx.doi.org/10.15154/z563-zd24. DOIs can be found at https://nda.nih.gov/abcd/abcd-annual-releases.

## Author Contributions

Conceptualization – EYX, ASFK, SML, HCW

Methodology: EYX, ASFK, SML, HCW

Formal analysis: EYX, PZG, ASFK

Data Curation: EYX, PZG

Writing – Original Draft: EYX

Writing – Review & Editing: All authors.

Supervision: ASFK, SML, HCW

