## Supplementary Information for "Longitudinal Prediction of Adolescent Depression from Environmental and Polygenic Risk Scores"

### Supervised machine learning

Supervised machine learning methods are widely used in developing risk prediction models. Broadly speaking, the goal of such models is to predict a given output (e.g. depression) based on a pre-selected set of input variables (also referred to as "predictors" or "features")[1,2]. Algorithms such as Elastic Net can predict continuous outcomes, handle large numbers of multicollinear variables, and perform feature selection – reducing a large feature set by shrinking coefficients of less relevant features[3–5]. This results in a simpler model which has a lower chance of overfitting errors – where a model matches the training data so closely that it cannot make accurate predictions in an independent sample[3,6]. Simpler models are also easier to implement both clinically and in independent samples, as predictor variables can be time-consuming and costly to measure – especially if these involve neuroimaging or biosample data.

The Elastic Net algorithm combines L1 (Least absolute shrinkage and selection operator; LASSO) and L2 (ridge) penalties in a regularized linear regression, allowing feature coefficients to be shrunk towards zero (ridge) or removed entirely (LASSO)[3–5]. The hyperparameter $\alpha$ (range 0-1) defines the degree of mixing between L1 and L2 penalties, where $\alpha = 0$ is equivalent to ridge regression and $\alpha=1$ is equivalent to LASSO. The hyperparameter $\lambda$ controls the strength of shrinkage and takes values from 0 to $+\infty$.[7]

Elastic Net models were fit in R version 4.4.3[8] using the 'glmnet' package version 4.1.8[9]. We used 10-fold cross-validation to tune both $\alpha$ and $\lambda$ hyperparameters, randomly dividing the training data into 10 folds. We tested 10 values for $\alpha$, increasing from 0 (ridge) to 1 (LASSO) in increments of 0.1 over 10 Elastic Net models. For each $\alpha$ value, 9 folds (i.e. 90%) were used to tune $\lambda$ which was then applied to the remaining fold[9,10]. After this was repeated across all 10 folds and for each $\alpha$, the result with $\alpha$ and $\lambda$ values that minimised mean squared-error was selected as the best-fitting model.

### Sensitivity analyses

#### 1. Depression score prediction using sex assigned at birth (Sensitivity analysis 1)

Elastic net models estimating 2-year depression scores and concurrent depression scores were re-trained using sex assigned at birth in place of gender as a predictor; the remaining features were unchanged (Table S3).

The best-fitting model predicting 2-year depression scores retained all 23 features, though the majority (16) of features had coefficients close to zero. 6 environmental factors had coefficients greater than 0.1 (Table S4). The best-fitting model was LASSO ($\alpha = 1$) with a test sample MSE = 0.84, explaining 16.5% variance in CBCL depression scores in the test sample. For depression scores measured concurrently with environmental factors, 8 factors had coefficients greater than 0.1 (Table S4). This model had $\alpha = 0.9$, with a test MSE = 0.81 and explained 19.1% of depression score variance in the test sample.

#### 2. Internalising score prediction using baseline features (Sensitivity analysis 2)

The same feature set was used in Elastic net models to predict CBCL internalising subscale scores measured at 2-year follow-up and concurrently at baseline (Table S3).

For CBCL internalising scores at 2-year follow-up, the best-fitting model was a ridge regression ($\alpha = 0$). This explained 18.6% of variance in internalising scores in the test sample with MSE = 0.82 in the test sample. 9 features had coefficients greater than 0.1 (Table S4) with the leading contributors being parental depression, emotional abuse and sexual abuse. For concurrent internalising scores, the best-fitting model was again a ridge regression (α = 0). This model explained 20.8% of variance in the test sample with a MSE = 0.79. 8 environmental factors had coefficients greater than 0.1 (Table S3) with leading contributors being parental depression, dieting and physical abuse.

#### 3. Cross-sectional prediction using 2-year follow-up features (Sensitivity analysis 3)

Demographic characteristics of participants included in this analysis are described in Table S5. 32 features were available at ABCD 2-year follow-up. Features unique to this time-point included bullying, substance use days, chronotype, dietary intake, unpredictable life events and effortful control (Table S3, Table S6). Using these environmental factors, we estimated CBCL DSM5-oriented depression and CBCL internalising subscale scores measured concurrently at 2-year follow-up.

The best-fitting cross-sectional model for depression scores was a ridge regression (α = 0), with MSE = 0.67 in the testing sample. These explained greater variance in test sample scores (32.7%) compared to the equivalent cross-sectional model at baseline (18.3%). For internalising scores, the best-fitting model was also ridge regression (α = 0), with MSE = 0.71 in the testing sample. Similarly, this feature set explained greater variance in test sample scores (29.4%) compared to the equivalent cross-sectional model at baseline (20.8%).

#### 4. depRS and European PRS predicting lifetime MDD (Sensitivity analysis 4)

PRS for participants of European ancestry were generated using European-only GWAS summary statistics[11]; the same diverse ancestry summary statistics were used to generate PRS for participants of African, American Admixed and East Asian ancestries. PRS-only prediction of lifetime MDD status at 2-year follow-up was better-than-chance and better classified parent-reported compared to youth self-reported lifetime MDD; using European-only summary statistics improved accuracy of parent-reported, but not youth-reported MDD (Table S7). The linear combination of PRS and depRS improved overall model fit for both outcomes compared to depRS-only prediction (Parent: $\chi^2$ = 25.79; p<.001; Youth: $\chi^2$ = 25.74; p<.001), with a slight (1%) increase in prediction accuracy. Including a depRS x PRS interaction maximised classification of parent-reported lifetime MDD status (AUC = 0.71 [95% CI 0.65 – 0.79]), but this term did not associate with lifetime MDD odds (Table S7).

**Table S1: Demographic characteristics of ABCD participants excluded from analyses**

| | (i) Baseline (N = 4292) | (ii) 2-year follow-up (N = 4832) | (iii) PRS youth (N = 5562) | (iv) PRS parent (N = 5602) |
|---|---|---|---|---|
| Age, mean (SD), years | | | | |
| Baseline | 9.92 (0.63) | 9.92 (0.63) | 9.92 (0.63) | 9.92 (0.63) |
| 2-year follow-up | NA | 12.03 (0.68) | 12.04 (0.67) | 12.04 (0.67) |
| Gender identity [1], n (%) | | | | |
| Cisgender male | 2189 (51.1%) | 2450 (50.79%) | 2813 (50.65%) | 2845 (50.86%) |
| Cisgender female | 2087 (48.72%) | 2366 (49.05%) | 2732 (49.19%) | 2740 (48.98%) |
| Transgender male | <5 (<0.1%) | <5 (<0.1%) | <5 (<0.1%) | <5 (<0.1%) |
| Transgender female | <5 (<0.1%) | <5 (<0.1%) | <5 (<0.1%) | <5 (<0.1%) |
| Gender non-conforming | 6 (0.1%) | 6 (0.12%) | 6 (0.11%) | 6 (0.11%) |
| Assigned sex at birth, n (%) | | | | |
| Male | 2191 (51.04%) | 2451 (50.71%) | 2815 (50.6%) | 2847 (50.81%) |
| Female | 2099 (48.89%) | 2379 (49.22%) | 2745 (49.34%) | 2753 (49.13%) |
| Intersex | <5 (<0.1%) | <5 (<0.1%) | <5 (<0.1%) | <5 (<0.1%) |
| Race/Ethnicity [2], n (%) | | | | |
| Asian | 86 (2%) | 101 (2.09%) | 191 (3.43%) | 191 (3.41%) |
| Black | 820 (19.11%) | 937 (19.4%) | 988 (17.77%) | 998 (17.82%) |
| Hispanic | 1003 (23.37%) | 1135 (23.49%) | 1279 (23%) | 1289 (23.01%) |
| White | 1965 (45.79%) | 2179 (45.1%) | 2393 (43.03%) | 2407 (42.97%) |
| Other | 417 (9.72%) | 479 (9.92%) | 710 (12.77%) | 716 (12.78%) |
| Total household income p.a. [3], n (%) | | | | |

| | (i) Baseline (N = 4292) | (ii) 2-year follow-up (N = 4832) | (iii) PRS youth (N = 5562) | (iv) PRS parent (N = 5602) |
|---|---|---|---|---|
| < $5,000 | 178 (5.43%) | 215 (5.63%) | 242 (5.32%) | 245 (5.34%) |
| $5,000 - $11,999 | 143 (4.36%) | 186 (4.87%) | 212 (4.66%) | 215 (4.68%) |
| $12,000 - $15,999 | 95 (2.9%) | 121 (3.17%) | 139 (3.05%) | 140 (3.05%) |
| $16,000 - $24,999 | 193 (5.88%) | 230 (6.02%) | 266 (5.84%) | 268 (5.84%) |
| $25,000 - $34,999 | 211 (6.43%) | 250 (6.54%) | 292 (6.42%) | 293 (6.38%) |
| $35,000 - $49,999 | 283 (8.63%) | 341 (8.92%) | 389 (8.55%) | 396 (8.63%) |
| $50,000 - $74,999 | 433 (13.2%) | 517 (13.53%) | 594 (13.05%) | 598 (13.03%) |
| $75,000 - $99,999 | 443 (13.5%) | 489 (12.8%) | 595 (13.07%) | 600 (13.07%) |
| $100,000 - $199,999 | 942 (28.71%) | 1058 (27.69%) | 1283 (28.19%) | 1292 (28.14%) |
| <$200,000 | 360 (10.97%) | 414 (10.83%) | 539 (11.84%) | 544 (11.85%) |
| Parental education, n (%) | | | | |
| <High school | 447 (10.45%) | 502 (10.42%) | 549 (9.9%) | 555 (9.94%) |
| High school/GED | 542 (12.68%) | 619 (12.85%) | 683 (12.32%) | 687 (12.3%) |
| Some College/Associate degree | 1323 (30.94%) | 1517 (31.5%) | 1672 (30.15%) | 1686 (30.18%) |
| Bachelor’s degree | 1087 (25.42%) | 1197 (24.85%) | 1388 (25.03%) | 1394 (24.96%) |
| Postgraduate degree | 877 (20.51%) | 981 (20.37%) | 1254 (22.61%) | 1264 (22.63%) |
| Lifetime MDD, n (%) | | | | |
| 0 | NA | NA | 2547 (95.86%) | 2488 (95.84%) |
| 1 | NA | NA | 110 (4.14%) | 108 (4.16%) |
| Ancestry, n (%) | | | | |
| African | NA | NA | 1052 (24.25%) | 1063 (24.28%) |
| American Admixed | NA | NA | 1143 (26.35%) | 1156 (26.4%) |
| East Asian | NA | NA | 34 (0.78%) | 34 (0.78%) |
| European | NA | NA | 2109 (48.62%) | 2125 (48.54%) |

Table S1: Sample sizes and demographic characteristics for ABCD participants excluded from Elastic Net models predicting (i) concurrent and (ii) 2-year follow-up depression; and for participants excluded from depRS and PRS prediction of (iii) youth self-reported and (iv) parent-reported lifetime MDD at 2-year follow-up. Due to missingness in demographic data, numbers do not sum to the total sample size.

PRS = polygenic risk score; NA = not applicable in this sample; GED = General Educational Development diploma; MDD = major depressive disorder.

[1] To protect the privacy of study participants, demographic groups with fewer than 5 participants are denoted as <5 (<0.1%).

[2] The 5-level race/ethnicity variable was coded in ABCD Releases from caregiver responses to "What race do you consider the child to be? Please check all that apply." and "Do you consider the child Hispanic/Latino/Latina? (Yes/No)". "Other race/ethnicity" consists of non-Hispanic youth where "Other race" or multiple of the following races/ethnicities were selected: White, Black/African American, Alaska Native, Native Hawaiian, Guamanian, Samoan, Other Pacific Islander, Asian Indian, Chinese, Filipino, Japanese, Korean, Vietnamese, Other Asian, Other Race, Refuse to Answer, Don't Know.

[3] Total income before taxes or deductions from all sources – e.g. wages, benefits, rent from properties, social security, child payments etc.

**Table S2: Baseline demographic characteristics of participants included in PRS analyses by lifetime MDD status**

| | (i) Parent-reported | | (ii) Youth self-reported | |
|---|---|---|---|---|
| | **Lifetime MDD (N = 248)** | **No Lifetime MDD (N = 6011)** | **Lifetime MDD (N = 211)** | **No Lifetime MDD (N = 6088)** |
| Age, mean (SD), years | | | | |
| Baseline | 10 (0.63) | 9.9 (0.62) | 9.95 (0.62) | 9.91 (0.62) |
| 2-year follow-up | 12.15 (0.7) | 12.01 (0.66) | 12.15 (0.7) | 12.01 (0.66) |
| Gender identity [1], n (%) | | | | |
| Cisgender male | 123 (49.6%) | 3211 (53.42%) | 82 (38.86%) | 3284 (53.94%) |
| Cisgender female | 124 (50%) | 2798 (46.55%) | 129 (61.14%) | 2801 (46.01%) |
| Transgender male | <5 (<2.0%) | 0 (0%) | 0 (0%) | <5 (<0.1%) |
| Transgender female | 0 (0.0%) | <5 (<0.1%) | 0 (0%) | <5 (<0.1%) |
| Assigned sex at birth, n (%) | | | | |
| Male | 124 (50%) | 3214 (53.47%) | 81 (38.39%) | 3289 (54.02%) |
| Female | 124 (50%) | 2797 (46.53%) | 130 (61.61%) | 2799 (45.98%) |
| Race/Ethnicity [2], n (%) | | | | |
| Asian | <5 (<2.0%) | 58 (0.96%) | <5 (<2.0%) | 59 (0.97%) |
| Black | 28 (11.29%) | 756 (12.58%) | 41 (19.43%) | 753 (12.37%) |
| Hispanic | 45 (18.15%) | 1074 (17.87%) | 49 (23.22%) | 1080 (17.74%) |
| White | 141 (56.85%) | 3623 (60.27%) | 95 (45.02%) | 3683 (60.5%) |
| Other | 31 (12.5%) | 500 (8.32%) | 24 (11.37%) | 513 (8.43%) |
| Total household income p.a. [3], n (%) | | | | |
| < $5,000 | 7 (2.82%) | 165 (2.74%) | 13 (6.16%) | 162 (2.66%) |
| $5,000 - $11,999 | 7 (2.82%) | 200 (3.33%) | 8 (3.79%) | 202 (3.32%) |
| $12,000 - $15,999 | 6 (2.42%) | 127 (2.11%) | 6 (2.84%) | 128 (2.1%) |

| | (i) Parent-reported | | (ii) Youth self-reported | |
|---|---|---|---|---|
| | **Lifetime MDD (N = 248)** | **No Lifetime MDD (N = 6011)** | **Lifetime MDD (N = 211)** | **No Lifetime MDD (N = 6088)** |
| $16,000 - $24,999 | 15 (6.05%) | 240 (3.99%) | 19 (9%) | 238 (3.91%) |
| $25,000 - $34,999 | 17 (6.85%) | 344 (5.72%) | 13 (6.16%) | 349 (5.73%) |
| $35,000 - $49,999 | 31 (12.5%) | 507 (8.43%) | 22 (10.43%) | 523 (8.59%) |
| $50,000 - $74,999 | 34 (13.71%) | 866 (14.41%) | 53 (25.12%) | 851 (13.98%) |
| $75,000 - $99,999 | 41 (16.53%) | 928 (15.44%) | 26 (12.32%) | 948 (15.57%) |
| $100,000 - $199,999 | 72 (29.03%) | 1946 (32.37%) | 35 (16.59%) | 1992 (32.72%) |
| <$200,000 | 18 (7.26%) | 688 (11.45%) | 16 (7.58%) | 695 (11.42%) |
| Parental education, n (%) | | | | |
| <High school | 5 (2.02%) | 224 (3.73%) | 17 (8.06%) | 218 (3.58%) |
| High school/GED | 18 (7.26%) | 555 (9.23%) | 26 (12.32%) | 551 (9.05%) |
| Some College/Associate degree | 84 (33.87%) | 1710 (28.45%) | 79 (37.44%) | 1729 (28.4%) |
| Bachelor's degree | 72 (29.03%) | 1863 (30.99%) | 53 (25.12%) | 1888 (31.01%) |
| Postgraduate degree | 69 (27.82%) | 1659 (27.6%) | 36 (17.06%) | 1702 (27.96%) |
| Ancestry, n (%) | | | | |
| African | 36 (14.52%) | 972 (16.17%) | 57 (27.01%) | 962 (15.8%) |
| American Admixed | 58 (23.39%) | 1306 (21.73%) | 55 (26.07%) | 1322 (21.71%) |
| East Asian | <5 (<2.0%) | 66 (1.1%) | <5 (<2.0%) | 67 (1.1%) |
| European | 151 (60.89%) | 3667 (61%) | 97 (45.97%) | 3737 (61.38%) |

Table S2: Sample sizes and demographic characteristics by lifetime MDD status for ABCD participants included in depRS and PRS prediction of (i) youth self-reported and (ii) parent-reported lifetime MDD at 2-year follow-up.

PRS = polygenic risk score; NA = not applicable in this sample; GED = General Educational Development diploma; MDD = major depressive disorder.

[1] To protect the privacy of study participants, demographic groups with fewer than 5 participants are denoted as <5; numbers will sum to greater than the total sample size.

[2] The 5-level race/ethnicity variable was coded in ABCD Releases from caregiver responses to "What race do you consider the child to be? Please check all that apply." and "Do you consider the child Hispanic/Latino/Latina? (Yes/No)". "Other race/ethnicity" consists of non-Hispanic youth where "Other race" or multiple of the following races/ethnicities were selected: White, Black/African American, Alaska Native, Native Hawaiian, Guamanian, Samoan, Other Pacific Islander, Asian Indian, Chinese, Filipino, Japanese, Korean, Vietnamese, Other Asian, Other Race, Refuse to Answer, Don't Know.

[3] Total income before taxes or deductions from all sources – e.g. wages, benefits, rent from properties, social security, child payments etc.

**Table S3: Risk factors included in depRS**

| Risk factor | ABCD Variable Name(s) | Description | At baseline | At 2-year follow-up |
|---|---|---|---|---|
| Age (months) | interview_age | Age at assessment | Yes | Yes |
| Area deprivation | reshist_addr1_adi_perc | Area deprivation index national percentile for primary address. Higher values = greater deprivation. | Yes | Yes |
| BMI [1] | Height: anthro_1_height_in, anthro2heightin, anthro3heightin; Weight: anthroweight1lb, anthroweight2lb, anthroweight3lb) | Mean weight (lbs) /mean height (in) * 703. | Yes | Yes |
| Community safety | nsc_p_ss_mean_3_items | Neighborhood Safety/Crime Survey mean summary score. Lower values = less safe from crime. | Yes | Yes |
| Days of physical activity [2] | physical_activity1_y | Days of physical activity (active for at least 60 minutes) in the past week. | Yes | Yes |
| Loss of a loved one | ksads_ptsd_raw_770_p | “Learned about the sudden unexpected death of a loved one”. 0 = No; 1 = Yes. | Yes | Yes |
| Emotional abuse | ksads_ptsd_raw_764_p, ksads_ptsd_raw_765_p | Any of: “A non-family member threatened to kill your child”, “A family member threatened to kill your child”. 0 = No; 1 = Yes. | Yes | Yes |
| Family conflict [2] | fes_y_ss_fc_pr | FES Conflict subscale summary score. Higher values = greater conflict. | Yes | Yes |
| Gender (*sex in sensitivity analysis 1*) | demo_gender_id_v2 *(demo_sex_v2)* | M = cisgender and transgender males; F = cisgender and transgender females. *(M =* | Yes | Yes |

| Risk factor | ABCD Variable Name(s) | Description | At baseline | At 2-year follow-up |
|---|---|---|---|---|
| | | *assigned male at birth; F = assigned female at birth)*. | | |
| Household income | demo_comb_income_v2, demo_comb_income_v2_l | Total combined family income for the past 12 months.<br>1 = Less than $5,000 to 10 = $200,000 and greater. | Yes | Yes |
| Needed food but could not afford it | demo_fam_exp1_v2; demo_fam_exp1_v2_l | "In the past 12 months, has there been a time when you and your immediate family experienced any of the following: Needed food but couldn't afford to buy it or couldn't afford to go out to get it?"<br>0 = No; 1 = Yes. | Yes | Yes |
| Physical abuse | ksads_ptsd_raw_761_p, ksads_ptsd_raw_762_p, ksads_ptsd_raw_763_p | Any of: "Shot, stabbed, or beaten brutally by a non-family member", "Shot, stabbed, or beaten brutally by a grown up in the home", "Beaten to the point of having bruises by a grown up in the home".<br>0 = No; 1 = Yes. | Yes | Yes |
| Parental acceptance [2] | crpbi_y_ss_parent | CRPBI Acceptance subscale. Mean summary score for primary caregiver.<br>Lower values = lower acceptance. | Yes | No |
| Parental depression | asr_scr_depress_r | ASR Depressive Problems DSM-5-oriented scale raw score.<br>Higher values = greater depression. | Yes | Yes |
| Parental monitoring [2] | pmq_y_ss_mean | PMQ Parental Monitoring mean summary score.<br>Lower values = less monitoring. | Yes | Yes |
| Parental education level | demo_prnt_ed_v2, demo_prnt_ed_v2_2yr_l, demo_prnt_ed_v2_l | "What is the highest grade or level of school you have completed or the highest degree you have received?"<br>1 = <HS; 2 = HS/GED; 3 = Some college or | Yes | Yes |

| Risk factor | ABCD Variable Name(s) | Description | At baseline | At 2-year follow-up |
|---|---|---|---|---|
| | | Associate degree; 4 = Bachelor's degree; 5 = Postgraduate degree. | | |
| Sexual abuse | ksads_ptsd_raw_767_p, ksads_ptsd_raw_768_p, ksads_ptsd_raw_769_p | Any of :"A grown up in the home touched your child in their privates, had your child touch their privates, or did other sexual things to your child", "An adult outside your family touched your child in their privates, had your child touch their privates or did other sexual things to your child", "A peer forced your child to do something sexually".<br>0 = No; 1 = Yes. | Yes | Yes |
| Was in a serious accident | ksads_ptsd_raw_754_p, ksads_ptsd_raw_755_p | Any of: "A car accident in which your child or another person in the car was hurt bad enough to require medical attention", "Another significant accident for which your child needed specialized and intensive medical treatment".<br>0 = No; 1 = Yes. | Yes | Yes |
| Hours of sleep | sleepdisturb1_p | "How many hours of sleep does your child get on most nights?" in the past 6 months (reverse coded).<br>1 = <5h; 2 = 5-7h; 3 = 7-8h; 4 = 8-9h; 5 = 9h+ | Yes | Yes |
| Tried tobacco [2,3] | tlfb_tob_puff | "Have you ever tried a puff from a tobacco or electronic cigarette, or vape pens, or e-hookah at any time in your life?".<br>0 = No; 1 = Yes. | Yes | No |
| Dieting to lose weight | ksads_13_72_p | KSADS Present symptom – weight control other (laxatives, exercise, dieting, pills).<br>0 = No; 1 = Yes. | Yes | Yes |

| Risk factor | ABCD Variable Name(s) | Description | At baseline | At 2-year follow-up |
|---|---|---|---|---|
| Witnessed violence in the community | ksads_ptsd_raw_760_p | "Witnessed someone shot or stabbed in the community".<br>0 = No; 1 = Yes. | Yes | Yes |
| Witnessed domestic violence | ksads_ptsd_raw_766_p | "Witness the grownups in the home push, shove or hit one another".<br>0 = No; 1 = Yes. | Yes | Yes |
| Bullying victimisation [2] | peq_ss_relational_victim, peq_ss_reputation_victim, peq_ss_overt_victim | Sum of PEQ Relational, Reputational and Overt victimization subscales.<br>Higher values = greater frequency of peer victimization. | No | Yes |
| Chronotype [2] | mctq_msfsc_calc | MCTQ chronotype (i.e. corrected local time of mid-sleep on non-school days). | No | Yes |
| Cyberbullying [2] | cybb_phenx_harm | "Have you ever been cyberbullied, where someone was trying on purpose to harm you or be mean to you online, in texts, or group texts, or on social media (like Instagram or Snapchat)?"<br>0 = No; 1 = Yes. | No | Yes |
| Discrimination [2] | dim_y_ss_mean | Experiences of being treated unfairly or negatively due to ethnic group, mean summary score of values from:<br>1 = Almost never to 5 = Very often. | No | Yes |
| Uncertain life events | ple_p_ss_total_number | Total number of stressful life events during the past 12 months which child had little/no control over, e.g. parent figure losing job, moving home, close friend was seriously ill/injured. | No | Yes |
| Effortful control | eatq_p_ss_effort_cont_ss | EATQ Effortful Control composite scale from Attention, Activation Control and Inhibitory Control subscales.<br>Lower values = lower self-regulation. | No | Yes |

| Risk factor | ABCD Variable Name(s) | Description | At baseline | At 2-year follow-up |
|---|---|---|---|---|
| Healthy diet: fiber [4] | bkfs_dt_fibe | BKFS estimated daily fiber intake in grams. | No | Yes |
| Healthy diet: fruits | bkfs_fruit_ce | BKFS average daily fruit/fruit juice intake in cup equivalents. | No | Yes |
| Healthy diet: vegetables | bkfs_vegnopot_ce | BKFS average daily vegetable intake in cup equivalents. | No | Yes |
| Substance(s) use days [2] | tlfb_cal_scr_num_events | TLFB number of recorded substance use events. Multiple usages per day for a substance are pooled together. | No | Yes |

Table S3: Features included in depRS measured at ABCD baseline and additional features at 2-year follow-up (sensitivity analysis 3). All variables were parent-reported unless indicated otherwise in bold text. ABCD = Adolescent Brain Cognitive Development study; FES = Family Environment Scale; CRPBI = Children's Report of Parental Behavior Inventory; ASR = Adult Self-Report; DSM-5 = Diagnostic and Statistical Manual of Mental Disorders, 5th Edition; PMQ = Parental Monitoring Questionnaire; HS = High School; GED = General Educational Development test; KSADS = Kiddie Schedule for Affective Disorders and Schizophrenia; PEQ = Peer Experiences Questionnaire; MCTQ = Munich Chronotype Questionnaire; EATQ = Early Adolescent Temperament Questionnaire; BKFS = Block Kids Food Screener; TLFB = Time Line Follow Back.

[1] Mean height and weight calculations were available in ABCD, however these were re-calculated in the present analyses due to data entry errors which yielded impossible values for height and weight (e.g. height of 257in (653 cm) and weight of 6201 lbs (2,813kg)). Following ABCD protocol, mean height and weight was calculated from the two closest measurements where available. Modified z-scores were calculated based on 2022 CDC extended BMI-for-age growth charts (https://www.cdc.gov/growthcharts/extended-bmi.htm); biologically implausible values (z-scores <-5 and >+8) were then removed.

For additional details see Wei R et al. (2020) A method for calculating BMI z-scores and percentiles above the 95th percentile of the CDC growth charts. Ann Hum Biol. 2020;47(6):514-521. https://doi.org/10.1080/03014460.2020.1808065 and Freedman DS et al. (2015) Validity of the WHO cutoffs for biologically implausible values of weight, height, and BMI in children and adolescents in NHANES from 1999 through 2012. Am J Clin Nutr. 2015;102(5):1000. https://doi.org/10.3945/AJCN.115.115576

[2] Denotes a youth self-reported variable.

[3] Only administered if youth had heard of the substance.

[4] See Hunsberger, M. et al. (2015) Relative validation of Block Kids Food Screener. Matern Child Nutr. 2015;11: 260-270. https://doi.org/10.1111/j.1740-8709.2012.00446.x and Nagata, J.M et al. (2025) Social epidemiology of early adolescent nutrition. Pediatr Res. https://doi.org/10.1038/s41390-025-03838-z

**Table S4: depRS Elastic Net coefficients for features measured at baseline**

| | CBCL Depression | | | | Sensitivity 2: CBCL Internalising | |
|---|---|---|---|---|---|---|
| **Feature** | **Y2** | **Baseline** | **Sensitivity 1: Y2** | **Sensitivity 1: Baseline** | **Y2** | **Baseline** |
| Intercept | -0.30 | -1.08 | -0.46 | -0.99 | -0.82 | -1.5 |
| Age (months) | 0.04 | 0.02 | 0.04 | 0.03 | 0.01 | 0.02 |
| Area deprivation | -0.03 | -0.08 | -0.03 | -0.09 | -0.04 | -0.06 |
| BMI | 0.01 | -0.02 | 0.02 | -0.02 | 0 | 0 |
| Community safety | -0.02 | -0.05 | 0.02 | -0.05 | -0.04 | -0.06 |
| Days of physical activity | -0.01 | -0.02 | -0.02 | -0.02 | -0.01 | -0.01 |
| Loss of a loved one | 0.04 | 0.07 | -0.02 | 0.07 | 0.1 | 0.11 |
| Emotional abuse | 0.21 | -0.02 | 0.06 | 0.04 | 0.25 | 0.07 |
| Family conflict | 0.02 | 0.03 | 0.27 | 0.04 | 0 | 0.03 |
| Gender (*sex in sensitivity analyses*) | -0.04 | 0.03 | 0.02 | 0.02 | -0.1 | -0.05 |
| Household income | 0.02 | 0.00 | 0.02 | 0.01 | 0.01 | 0 |
| Needed food but could not afford it | 0.00 | 0.05 | 0.01 | 0.05 | -0.01 | 0.04 |
| Physical abuse | 0.24 | 0.52 | 0.13 | 0.48 | 0.22 | 0.35 |
| Parental acceptance | -0.03 | -0.03 | -0.03 | -0.03 | -0.02 | -0.02 |
| Parental depression | 0.30 | 0.36 | 0.30 | 0.37 | 0.32 | 0.41 |
| Parental monitoring | -0.04 | -0.04 | -0.04 | -0.04 | -0.03 | -0.05 |
| Parental education level | 0.00 | -0.01 | -0.01 | -0.01 | 0 | 0 |
| Sexual abuse | 0.07 | 0.17 | 0.09 | 0.14 | 0.22 | 0.15 |
| Was in a serious accident | 0.07 | 0.11 | 0.08 | 0.13 | 0.1 | 0.16 |
| Hours of sleep | -0.08 | -0.18 | -0.07 | -0.19 | -0.01 | -0.06 |
| Tried tobacco | -0.07 | 0.14 | -0.06 | 0.07 | -0.03 | 0.32 |
| Dieting to lose weight | 0.12 | 0.41 | 0.12 | 0.44 | 0.16 | 0.39 |
| Witnessed violence in the community | -0.18 | 0.18 | -0.22 | 0.13 | -0.21 | 0.02 |
| Witnessed domestic violence | 0.17 | 0.19 | 0.21 | 0.20 | 0.17 | 0.2 |

Table S4: Standardised β coefficients for environmental factors predicting CBCL DSM5-oriented Depression and CBCL Internalising subscale scores at 2-year follow-up (Y2) and scores measured concurrently with environmental factors at ABCD baseline. Sensitivity analysis 1 used sex assigned at birth in place of gender identity as a predictor; sensitivity analysis 2 used the same baseline feature set to predict CBCL internalising problem scores.

**Table S5: Demographic characteristics for participants at 2-year follow-up (Sensitivity analysis 3)**

| | **2-year follow-up (N = 5,415)** |
|---|---|
| Age, mean (SD), years | 12 (0.6) |
| Gender identity [1], n (%) | |
| Cisgender male | 2,915 (53.8%) |
| Cisgender female | 2,489 (46.0%) |
| Transgender male | 7 (0.1%) |
| Transgender female | <5 (<0.1%) |
| Assigned sex at birth | |
| Male | 2,928 (54.1%) |
| Female | 2,487 (45.9%) |
| Race/Ethnicity [2], n (%) | |
| Asian | 125 (2.3%) |
| Black | 605 (11.2%) |
| Hispanic | 1,019 (18.8%) |
| White | 3,127 (57.7%) |
| Other | 539 (10.0%) |
| Household income p.a. [3], n (%) | |
| < $5,000 | 140 (2.6%) |
| $5,000 - $11,999 | 134 (2.5%) |
| $12,000 - $15,999 | 117 (2.2%) |
| $16,000 - $24,999 | 187 (3.5%) |
| $25,000 - $34,999 | 293 (5.4%) |
| $35,000 - $49,999 | 392 (7.2%) |
| $50,000 - $74,999 | 720 (13.3%) |
| $75,000 - $99,999 | 760 (14.0%) |
| $100,000 - $199,999 | 1,882 (34.8%) |
| <$200,000 | 790 (14.6%) |
| Parental education, n (%) | |
| <High school | 246 (4.5%) |
| High school/GED | 469 (8.7%) |
| Some College/Associate degree | 1,457 (26.9%) |
| Bachelor's degree | 1,625 (30.0%) |
| Postgraduate degree | 1,618 (29.9%) |

Table S5: Demographic characteristics for the sample of ABCD participants with complete data on environmental factors measured at 2-year follow-up. NA = not applicable in this sample; GED = General Educational Development diploma.

[1] To protect the privacy of study participants, demographic groups with fewer than 5 participants are denoted as <5 (<0.1%); numbers will sum to greater than the total sample size.

[2] The 5-level race/ethnicity variable was coded in ABCD Releases from caregiver responses to "What race do you consider the child to be? Please check all that apply." and "Do you consider

the child Hispanic/Latino/Latina? (Yes/No)". "Other race/ethnicity" consists of non-Hispanic youth where "Other race" or multiple of the following races/ethnicities were selected: White, Black/African American, Alaska Native, Native Hawaiian, Guamanian, Samoan, Other Pacific Islander, Asian Indian, Chinese, Filipino, Japanese, Korean, Vietnamese, Other Asian, Other Race, Refuse to Answer, Don't Know.

[3] Total income before taxes or deductions from all sources – e.g. wages, benefits, rent from properties, social security, child payments etc.

**Table S6: Elastic Net coefficients for features measured at 2-year follow-up (Sensitivity analysis 3)**

| Feature | CBCL Depression | CBCL Internalising |
|---|---|---|
| Intercept | -0.42 | -0.89 |
| Age (months) | 0.03 | 0.01 |
| Area deprivation | -0.05 | -0.05 |
| Dietary fiber intake | 0.03 | 0.03 |
| Fruit intake | -0.03 | -0.01 |
| VegTable S intake | -0.04 | -0.02 |
| BMI | 0.01 | 0.02 |
| Bullying * Gender | 0.00 | -0.02 |
| Bullying victimisation | 0.09 | 0.10 |
| Chronotype | 0.01 | 0.00 |
| Community safety | 0.00 | -0.02 |
| Cyberbullying | 0.07 | 0.04 |
| Days of physical activity | -0.02 | -0.02 |
| Loss of a loved one | -0.01 | 0.03 |
| Discrimination | -0.01 | -0.01 |
| Effortful control | -0.22 | -0.18 |
| Emotional abuse | 0.22 | 0.11 |
| Family conflict | 0.01 | -0.01 |
| Gender | -0.03 | -0.06 |
| Household income | 0.01 | 0.00 |
| Uncertain life events | 0.11 | 0.14 |
| Needed food but could not afford it | 0.01 | 0.00 |
| Physical abuse | 0.24 | 0.26 |
| Parental depression | 0.26 | 0.31 |
| Parental monitoring | -0.05 | -0.03 |
| Parental education level | -0.01 | 0.00 |
| Sexual abuse | 0.28 | 0.48 |
| Was in a serious accident | 0.09 | 0.09 |
| Hours of sleep | -0.10 | -0.01 |
| Substance use days | 0.02 | 0.01 |
| Dieting to lose weight | 0.35 | 0.28 |

Table S6: Standardised β coefficients for environmental factors associated with concurrent CBCL DSM5-oriented Depression and CBCL Internalising subscale scores measured at 2-year follow-up assessments.

**Table S7: depRS and PRS prediction of lifetime MDD using European-only summary statistics (Sensitivity analysis 4)**

| | **Model fit** | | | **Odds ratio [95% CI]** | | |
|---|---|---|---|---|---|---|
| **Model** | **AUC [95% CI]** | **$R^2$** | **RMSE** | **depRS [95% CI]** | **PRS [95% CI]** | **depRS * PRS [95% CI]** |
| Youth self-reported lifetime MDD | | | | | | |
| depRS | 0.64 [0.59 – 0.68] | 0.010 | 0.179 | 1.53 [1.37 – 1.71]*** | NA | NA |
| PRS | 0.59 [0.51 – 0.68] | 0.003 | 0.180 | NA | 1.33 [1.15 – 1.52]*** | NA |
| depRS + PRS | 0.65 [0.60 – 0.70] | 0.012 | 0.179 | 1.51 [1.35 – 1.68]*** | 1.26 [1.10 – 1.45]** | NA |
| depRS * PRS | 0.65 [0.62 – 0.67] | 0.013 | 0.179 | 1.50 [1.33 – 1.68]*** | 1.25 [1.08 – 1.46]** | 1.01 [0.91 – 1.13] |
| Parent-reported lifetime MDD | | | | | | |
| depRS | 0.68 [0.63 – 0.72] | 0.024 | 0.193 | 1.73 [1.57 – 1.91]*** | NA | NA |
| PRS | 0.61 [0.59 – 0.65] | 0.005 | 0.195 | NA | 1.44 [1.26 – 1.63]*** | NA |
| depRS + PRS | 0.69 [0.62 – 0.75] | 0.028 | 0.192 | 1.69 [1.53 – 1.87]*** | 1.34 [1.18 – 1.53]*** | NA |
| depRS * PRS | 0.71 [0.65 – 0.79] | 0.028 | 0.192 | 1.70 [1.53 – 1.88]*** | 1.35 [1.17 – 1.56]*** | 0.99 [0.90 – 1.10] |

Table S7: Sensitivity analysis using PRS derived from European-only GWAS summary statistics for participants with European ancestry; PRS for all other ancestries are unchanged. Model fit and odds ratios from logistic regression models predicting youth self-reported and parent-reported lifetime MDD assessed at 2-year follow-up.

AUC = Area Under the Receiver Operator Curve. RMSE = Root Mean Square Error.  MDD = Major Depressive Disorder. depRS = Linear predictor from Elastic Net model predicting depression scores at 2-year follow-up from baseline environmental factors. PRS = Polygenic Risk Score. NA = not estimated in this model.

*p<0.05, **p<0.01, ***p<0.001
